# Longitudinal Masked Representation Learning for Pulmonary Nodule Diagnosis from Language Embedded EHRs

**DOI:** 10.1101/2025.05.09.25327341

**Authors:** Thomas Z. Li, John M. Still, Lianrui Zuo, Yihao Liu, Aravind R. Krishnan, Kim L. Sandler, Fabien Maldonado, Thomas A. Lasko, Bennett A. Landman

## Abstract

Electronic health records (EHRs) are a rich source of clinical data, yet exploiting longitudinal signals for pulmonary nodule diagnosis remains challenging due to the administrative noise and high level of clinical abstraction present in these records. Because of this complexity, classification models are prone to overfitting when labeled data is scarce. This study explores masked representation learning (MRL) as a strategy to improve pulmonary nodule diagnosis by modeling longitudinal EHRs across multiple modalities: clinical conditions, procedures, and medications. We leverage a web-scale text embedding model to encode EHR event streams into semantically embedded sequences. We then pretrain a bidirectional transformer using MRL conditioned on time encodings on a large cohort of general pulmonary conditions from our home institution. Evaluation on a cohort of diagnosed pulmonary nodules demonstrates significant improvement in diagnosis accuracy with a model finetuned from MRL (0.781 AUC, 95% CI: [0.780, 0.782]) compared to a supervised model with the same architecture (0.768 AUC, 95% CI: [0.766, 0.770]) when integrating all three modalities. These findings suggest that language-embedded MRL can facilitate downstream clinical classification, offering potential advancements in the comprehensive analysis of longitudinal EHR modalities.

## 1. Introduction

Electronic health records (EHRs) capture most of the important touch points patients have with the health care system, which makes them the most comprehensive source of clinical data available. Modeling longitudinal signals from EHRs for lung cancer diagnosis is largely unexplored. Extracting useful signals from EHRs is challenging however many are obscured by the multitude of administrative tasks that healthcare practitioners perform daily. For example, a single lab test can trigger a cascade of additional tests, seemingly unrelated diagnostic codes from differential diagnoses, and medication prescriptions from empirical treatments that may not target the true disease process. Common modeling approaches, such as counting events over a period or analyzing variables at a single time point, can destroy the longitudinal structure essential for modeling complex relationships.

Masked representation learning (MRL) has been empirically shown to be a leading method for learning longitudinal features. It involves masking a portion of features and training a model to recover this portion. For data with strong temporal relationships (i.e. time series, sequential data), masking in the time domain naturally emerges to have models predict the present from the past. BEHRT [1] and Med-BERT [2] applied masked token prediction to structured EHR sequences, improving disease prediction. In unstructured data, ClinicalBERT [3] and BioBERT [4] utilized MRL to learn representations from clinical text.

Despite these advances, longitudinal EHR modeling for pulmonary nodule diagnosis remains underexplored. Existing frameworks primarily focus on general-purpose pretraining with a single modality (i.e. ICD codes) within a short time frame. Our work addresses this gap by conducting masked representation learning over multi-year time scales across three modalities: conditions, procedures, and medications. We leverage an open-source language embedding to jointly represent these modalities and train a model to predict randomly masked events conditioned on their time encodings. For pretraining, we selected a large cohort encompassing patients with a broad set of pulmonary conditions. Evaluating on a separate cohort of patients with indeterminate pulmonary nodules, we find that MRL improves pulmonary nodule diagnosis compared to a supervised counterpart with the same model architecture. These results preliminarily demonstrate the potential for language embedded MRL to enable lifetime longitudinal analysis of EHRs towards a clinically meaningful diagnosis.

## 2. Methods

### 2.1 Data

All data was collected from the Research Derivative [5] of Vanderbilt University Medical Center under IRB #140274. We pulled variables from three modalities, diagnostic codes, procedural codes, and medication codes, as event streams from 2000 to 2024. Here we denote an event as recorded variable of any modality with a corresponding timestamp in the EHR. International Classification of Diseases, 9^th^ revision (ICD-9) and ICD-10 diagnostic codes were mapped to the Systematized Nomenclature of Medicine Clinical Terms (SNOMED) [6] vocabulary, representing diagnoses and conditions. Meanwhile, all Current Procedural Terminology (CPT) codes were retreived as procedural codes and medications were pulled from the database as RxNorm vocabulary [7]. A unique identifier was assigned to each unique code such that two events with the same code would share this identifier. In addition, each unique code had a concept name following their respective vocabulary, typically resembling a taxonomy standardized descriptive phrase. As all the vocabularies employed in this study have a hierarchical taxonomy, the concept names varied in specificity. For example, “Systolic heart failure” came from a lower level SNOMED code that fell under the parent code “Heart failure”. Lastly, we pulled the birthdate, birth sex, and race of each patient. Following the IRB protocol, dates were shifted and patient identifiers were hashed for anonymization purposes.

We built a *discovery set* of 109,031 patients selected for having a broad range pulmonary diseases according to the SNOMED vocabulary. This was used for pretraining the masked representation model in a self-supervised manner. To evaluate the usefulness of the learned representations in predicting lung cancer, we identified an *SPN set* of 13069 patients with a solitary pulmonary nodule (SPN) code and no prior history of any cancer. Lung cancer cases were identified as patients with a billing code for any lung malignancy occurring 4-1095 days post SPN date and controls were those with no such code during the same period [8]. The SPN set was split into training (n=10530) and testing (n=2639) sets (Table 1). There was incomplete overlap between the discovery set and SPN training set, but the discovery set and SPN test set were made to be disjoint.

**Table 1.** Characteristics of Discovery and Solitary Pulmonary Nodule (SPN) Datasets.

| **Dataset** | **Discovery Set** | **SPN Training Set** | **SPN Test Set** |
| --- | --- | --- | --- |
| Patients | 109,031 | 10,530 | 2639 |
| Events per patient |  |  |  |
| Conditions | 160±197 | 127±215 | 122±192 |
| Procedures | 102±128 | 54.9±103 | 53±91 |
| Medications | 217±238 | 573±1323 | 560±1313 |
| Lung Cancer (%) | N/A | 614 (0.06%) | 153 (0.06%) |

### 2.2 Natural Language Preprocessing

We removed patients who had either negligible or excess engagement with the health care system, defined as having less than 10 events or being in the 95^th^ percentile of record length respectively. We did not remove patients with excess engagement in the SPN sets because there was the possibility that lung nodule management may present as a dense record. We designed a natural language template for each variable combining its modality (i.e. condition, procedure, or medication) with its concept ID and concept name in the following form.

> *[Modality] #[Concept ID]: [Concept name]*

Although it has no semantic value, the concept ID was included to help distinguish between semantically similar but unique concepts. Treating the natural language form of each variable as a sentence, we computed a vector embedding for each variable from the state-of-the-art text embedding model (TEM) dunzhang/stella_en_400M_v5 (https://huggingface.co/dunzhang/stella_en_400M_v5) [9,10]. This specific TEM was chosen for its small memory footprint and high ranking performance on the Massive Text Embedding Benchmark [11]. First, a SentencePiece tokenizer [12] transformed the sentence into a sequence of sub-words across the entire dataset. Then, the TEM encodes the sequence and computes the average to generate a single text embedding for the variable. In this manner we precomputed embeddings for all unique variables, enabling event embeddings to be retrieved efficiently via a cached mapping back to their corresponding variable embeddings.

### 2.3 Masked Representation Learning

Our self-supervised model first projected the preprocessed event stream (natural language transformed and tokenized) of each patient into the TEM’s pretrained embedding space in ℝ^1024^. The input event stream was collated such that the output of the TEM was another sequence of tokens such that each was an embedded event. *Token* from here on refers to a vector embedding of an event as opposed to the sub-word encodings from the SentencePiece tokenizer. This allowed us to add a time encoding [13] to each token corresponding with the event’s timestamp. This sequence formed the input to a bidirectional transformer that was optimized from random weights using BERT-style MRL (Figure 1) [14].

**Figure 1.**
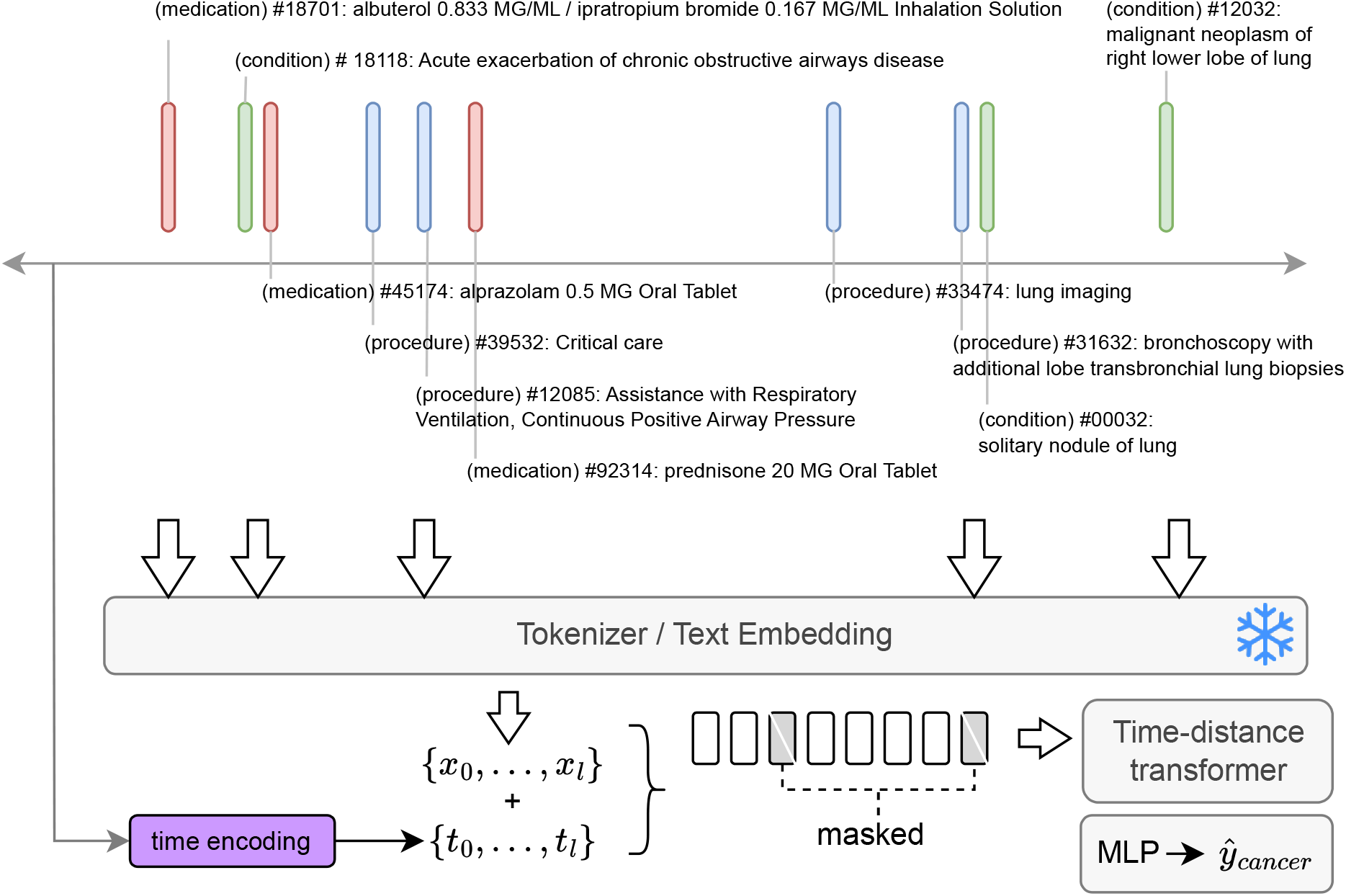
Streams of events (defined as an episodic variable and its timestamp) were pulled from the VUMC EHRs and preprocessed into natural language templates. We leveraged a leading text embedding model to embed each event as feature vectors {***x***_**0**_, …,***x***_***l***_}. Corresponding time encodings {***t***_**0**_, …,***t***_***l***_}. were added to each token corresponding to the event’s timestamp. From this sequence, we pretrained a transformer masked representation learning and finetuned on lung cancer labels.

During MRL, 15% of tokens are randomly and non-contiguously selected. Since the masking procedure is not done at the label-prediction stage, we do not mask all the selected tokens to prevent overfitting on the masked prediction task. Instead, only 80% of the initially selected tokens are replaced with a mask token, which is a vector embedding that is randomly initialized at the start of training and fixed during training. 10% of these signatures remain unchanged, and the remaining 10% are replaced with an “incorrect” variable selected at random from the corpus of possible variables. Importantly, time encodings were added to the masked tokens as well to provide the model with temporal context. Given the logits from the masked tokens, {*x*_1_, …,*x*_*N*_}, and a set of classes defined as the corpus of *m* unique variables {*c*_1_, …,*c*_*m*_}, the loss for a sequence (i.e. a single patient) is computed with cross-entropy as follows:

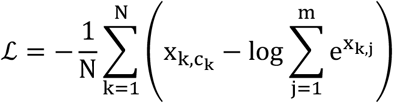

Where 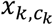 is the logit for the correct class of the *k^th^* token and *x*_*k*,*j*_ is the logit for class *j* in the *k^th^* token.

## 3. Experiments

We pretrained the masked representation model on the discovery set with the TEM module frozen. We evaluated the usefulness of this model (Table 1: MRL-finetune) through fine-tuned prediction of pulmonary nodule diagnosis within 3 years using the SPN training and test sets. During both pretrain and fine-tune stages we employed Adam optimization [15] and set the learning rate scheduler to cosine annealing with warm restart cycles [16]. We initialized finetuning with the pretrained MRL weights and added a “CLS” token [17], which fed into a multi-layer perceptron for classification. As a comparative baseline, we reused the model architecture in the fine-tune stage but trained from random weights (Table 1: Supervised). We repeated these experiments with a single modality in diagnostic codes and all three modalities.

Results are reported as mean area under the receiver operating characteristic curve (AUC) and 95% confidence intervals from 1000 bootstrap samples of the test set predictions, sampling with replacement. We used the two-sided Wilcoxon signed-rank test to determine if performance of each approach was significantly different than its supervised counterpart for the same set of modalities at *p*<0.05.

## 4. Results

When only given diagnostic codes, we did not observe a difference in pulmonary nodule diagnosis between MRL-finetune and the supervised models (0.773 [0.772, 0.774] vs, 0.772 [0.769, 0.775], resp.). In the tri-modality context, however, MRL-finetune (0.781 [0.780, 0.782]) significantly outperformed the supervised approach (0.768 [0.766, 0.770]) (Table 2).

**Table 2.**
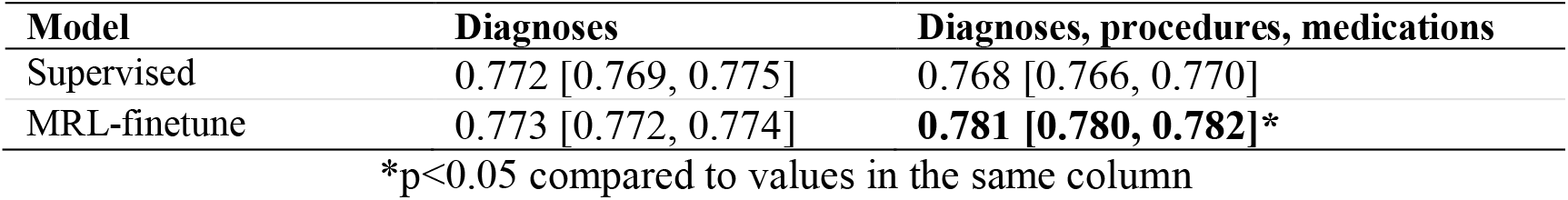
Mean AUC [95% CI] in single modality and tri-modality learning.

## 5. Discussion

This study is a preliminary investigation of enhancing three-year pulmonary nodule diagnosis using MRL on longitudinal EHRs spanning multiple decades. Our contributions are (1) the integration of time encodings with language embeddings developed from web-scale efforts as a basis for MRL and (2) leveraging this approach to improve the diagnosis of incidentally-detected pulmonary nodules from a purely supervised approach.

The nuances of health care administration specific to each modality, might explain why MRL only lead to performance improvements when all three modalities were jointly learned. The billing of diagnostic codes is known to be heavily abstracted from medical reality for many reasons including the fact that many initial diagnosis are incorrect but still recorded, lack of specificity from assigning parent-level codes when more specific codes would significantly alter the clinical context, and the time of billing being chronologically separated from the patient’s management [18–22]. We suspect that procedural codes suffer from a much smaller degree of abstraction as there is intuitively little ambiguity in the treatments that patients receive. However, the benefit of procedural codes is somewhat contradicted in the decreased performance of the supervised tri-modal model. We suspect that this model overfit on the expanded feature space introduced by the additional modalities [23,24]. As intended, our unsupervised MRL approach may have mitigated this overfitting, resulting in improved performance in the multi-modal experiment.

We also consider medications to be considerably abstracted in the EHR because elements such as dosage, units of measurement, route of administration, frequency, duration, and titration schedule are not seamlessly recorded. In this study, these elements were either included in the concept name (i.e. “acetaminophen 500 MG Oral Tablet”) or exceedingly difficult to capture without parsing clinical notes. In simplifying medication events to their concept names, we relied on masked representation learning to capture their temporal dynamics and semantic relationship with other modalities. In examining a t-SNE map of event streams embedded with the masked representation model from two patients, we observed clusters of multiple modalities suggesting overarching clinical concepts (Figure 2). Whether MRL effectively learns concepts of medication prescription is area of future investigation.

**Figure 2.**
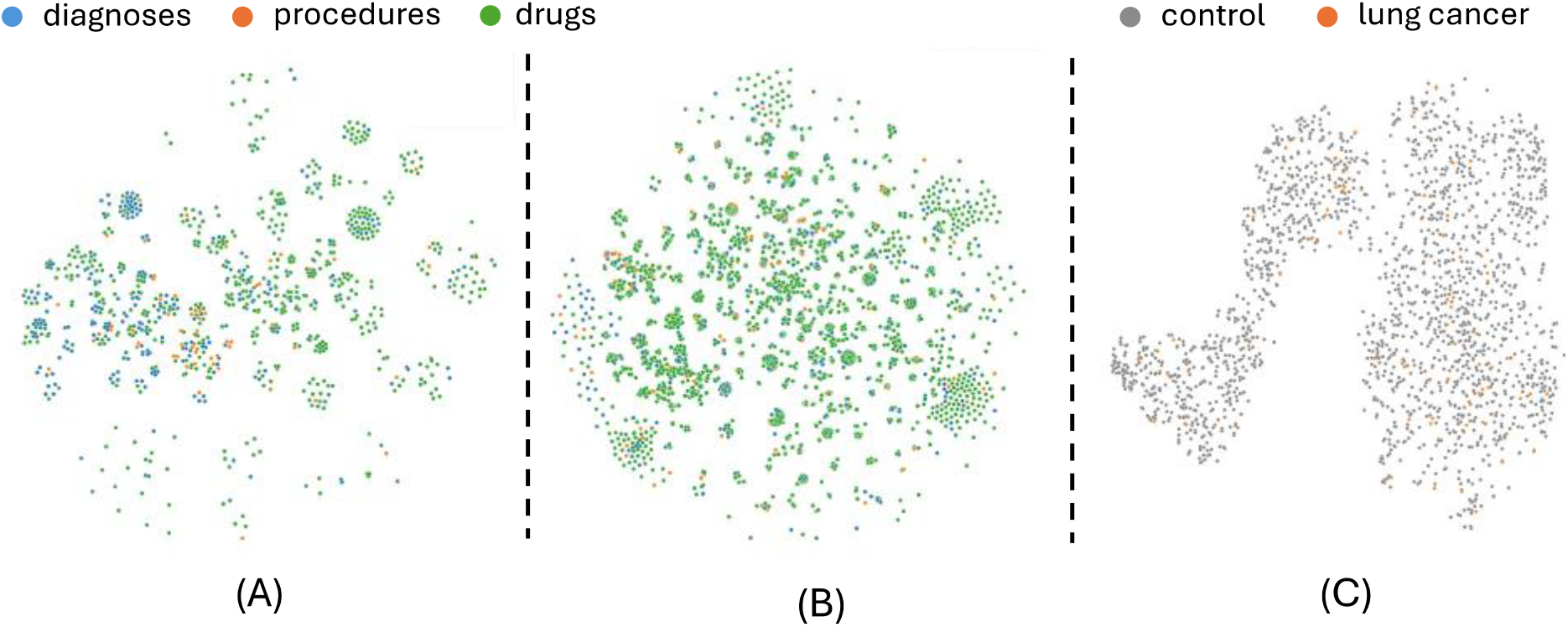
A. & B. Embeddings of event streams combining diagnoses, procedures and drugs together of two patients were visualized with t-SNE. Events from different modalities cluster together forming overarching clinical concepts. C. The embeddings of 30 events preceding a pulmonary nodule code were concatenated for 2000 patients and visualized with t-SNE.

## Data Availability

All data produced in the present study are available upon reasonable request to the authors

## 6. Acknowledgments

This research was funded by the NIH through F30CA275020, 2U01CA152662, and R01CA253923-02, as well as NSF CAREER 1452485 and NSF 2040462. This study was also funded by the Vanderbilt Institute for Surgery and Engineering through T32EB021937-07, the Vanderbilt Institute for Clinical and Translational Research through UL1TR002243-06, and the Pierre Massion Directorship in Pulmonary Medicine. We utilize generative AI to generate code segments based on task descriptions, as well as to assist with debugging, editing, and autocompleting code. Additionally, generative AI has been employed to refine sentence structure and ensure grammatical accuracy. However, all conceptualization, ideation, and prompts provided to the AI stem entirely from the authors’ creative and intellectual efforts. We take full responsibility for reviewing and verifying all AI-generated content in this work.

